# Re-evaluating progression and pathways following *Mycobacteria tuberculosis* infection within the spectrum of tuberculosis disease

**DOI:** 10.1101/2022.12.14.22283422

**Authors:** Katherine C. Horton, Alexandra S. Richards, Jon C. Emery, Hanif Esmail, Rein M. G. J. Houben

**Author notes:** KCH and ASR contributed equally to this work.

## Abstract

**Background:** Traditional understanding of the risk of progression from *Mycobacterium tuberculosis* (*Mtb*) infection to tuberculosis (TB) disease overlooks nuance across a spectrum of disease.

**Methods:** We developed a deterministic model of *Mtb* infection and minimal (pathological damage but not infectious), subclinical (infectious but no reported symptoms), and clinical (infectious and symptomatic) TB disease, informed by a rigorous evaluation of data from a systematic review of TB natural history. Using a Bayesian approach, we calibrated the model to data from historical cohorts that followed tuberculin-negative individuals to tuberculin conversion and TB disease, as well as data from cohorts that followed progression and regression between disease states, disease state prevalence ratios, disease duration, and mortality. We estimated incidence, pathways, and ten-year outcomes following *Mtb* infection for a simulated cohort.

**Results:** 90.8% (95% uncertainty interval, UI, 90.2-91.3) of individuals self-cleared within 10 years of infection, while 9.3% (95% UI 8.4-10.0) progressed to TB disease. Of those, 68.1% (95% UI 65.1-71.1) developed infectious disease, and 32.7% (95% UI 29.7-35.7) progressed to clinical disease. While 93% of progression to minimal disease occurred within two years of infection, only 63% and 38% of subclinical and clinical disease, respectively, occurred within this period. Multiple progression pathways from infection were necessary to calibrate the model, and 48.8% (95% UI 45.0-52.6) of those who developed infectious disease undulated between disease states.

**Conclusions:** We identified highly heterogeneous pathways across disease states after *Mtb* infection, highlighting the need for clearly defined disease thresholds to inform more effective prevention and treatment efforts to end TB.

## Introduction

Tuberculosis (TB) is a leading cause of morbidity and mortality and has substantial economic and social impacts worldwide (1). Despite growing acceptance that TB presents across a spectrum of disease states, defined by different pathological, bacteriological, and clinical thresholds (2-8), the pathways by which individuals progress from *Mycobacterium tuberculosis* (*Mtb*) infection and transition across thresholds are not well understood. Progression risks and timelines are founded instead on a conventional binary distinction between *Mtb* infection and infectious symptomatic TB disease. However, progression from infection to this advanced disease state is neither straightforward nor guaranteed.

*Mtb* infection is traditionally understood to confer a 5-10% lifetime risk of developing TB disease (1), with half of this risk occurring within two years of infection (9, 10). These axioms are typically understood to refer to infectious, usually symptomatic, disease (5, 11) and are often attributed to data from preventive chemotherapy trials (12) and Bacillus Calmette–Guérin (BCG) vaccination trials (13). However, timelines from these studies are, at best, approximate, as none documents the time at which individuals were infected with *Mtb*. These studies also reflect the binary approach that ignores nuance across the spectrum of disease and assumes unidirectional progression across a single threshold. Given the importance and wide reach of simplifying statements, the gaps in data for historical and more recent contributions to this canon are surprising.

Recognising these limitations, and in line with the shift towards understanding TB across a spectrum of disease, there is a clear need to re-evaluate our understanding of progression following *Mtb* infection. Intermediate disease states between *Mtb* infection and infectious symptomatic TB disease merit further attention. Infectious subclinical TB is as prevalent as infectious symptomatic disease globally (14) and likely contributes substantially to onward transmission (15-17). Pathological disease is also highly prevalent (18), and, even when non-infectious, can be severe. Focus on a single threshold of infectious symptomatic disease (19) ignores the potential contribution of intermediate disease states to morbidity and onward transmission and may be a key reason why progress to reduce TB incidence and mortality remains slower than needed to meet targets (20, 21).

Expanded efforts under the End TB Strategy (20) have rightly renewed focus on disease prevention, including amongst those infected with *Mtb* (20, 22, 23), with research and innovation priorities that include the development of safe and effective treatment regimens for *Mtb* infections and post-exposure prophylactic vaccines (24). Prevention strategies using these and other technologies are built on the binary approach, which informed the estimate of a quarter of the world carrying *Mtb* and at risk of progressing to TB disease (25), which is likely a substantial overestimate given new insights in the likelihood individuals clearing their *Mtb* infection (26, 27).

Development and implementation of novel prevention strategies, as well as resources for disease detection and treatment, require an updated understanding of the risk of progression following *Mtb* infection and subsequent pathways through the course of disease.

We therefore re-evaluate progression and pathways following *Mtb* infection across the spectrum of disease, examining different thresholds of disease based on pathological, bacteriological, and clinical characteristics, and allowing for self-clearance of Mtb infection. We build on recent work that quantified progression and regression between different TB disease states using an extensive systematic review of TB natural history (28) with mathematical modelling methods (29). We estimate incidence, pathways, and ten-year outcomes following *Mtb* infection across different disease thresholds.

## Methods

Our analysis examines pathways from *Mtb* infection through minimal, subclinical, and clinical TB disease states, with clearance from infection, recovery from minimal disease, and mortality from clinical disease. We focus solely on pulmonary TB in adults and adolescents.

### Definitions

<u>Infection</u>: Individuals with evidence of *Mtb* infection by an immunologic test in the absence of bacteriological evidence of TB disease or clinical signs or symptoms of TB disease (4, 5, 29, 30).

<u>Cleared</u>: Individuals who have effectively controlled or eliminated *Mtb* infection and will not progress to TB disease in the absence of reinfection (5, 6, 26).

<u>Recovered</u>: Individuals who have effectively controlled or eliminated *Mtb* infection after developing TB disease and will not progress to TB disease in the absence of reinfection.

<u>Minimal disease</u>: Individuals with inflammatory pathology prior to the onset of bacteriological evidence of TB disease or symptoms of active TB disease (29, 31).

<u>Subclinical disease</u>: Individuals with bacteriological evidence of TB disease who do not report symptoms of active TB disease on screening (14, 29).

<u>Clinical disease</u>: Individuals with bacteriological evidence of TB disease with symptoms of active TB disease (29).

<u>Infectious disease</u> refers to bacteriologically-positive disease; as such, it includes subclinical and clinical disease, but excludes minimal disease (29).

<u>TB disease</u>: Any state of minimal, subclinical, or clinical disease.

### Data synthesis

To synthesise evidence of progression following *Mtb* infection, we reviewed studies from a recent systematic review of TB natural history (28), as well as studies referenced by two recent papers on progression following *Mtb* infection (11, 32). We sought studies that followed cohorts of tuberculin-negative individuals to tuberculin conversion and then to TB disease (whether classified as minimal, subclinical or clinical). For inclusion, studies were required to document tuberculin conversion, report intervals between tuberculin testing, the number of individuals followed from tuberculin conversion for incident disease, intervals between disease screening following tuberculin conversion, the number of individuals who developed disease after tuberculin conversion, and the interval between tuberculin conversion and disease detection for those who developed disease.

We adjusted data to reflect uncertainty in the time of tuberculin conversion and disease onset, recognising that neither infection nor disease onset occurred at the point those developments were detected in these studies. To estimate time of infection, we sampled from a uniform distribution over each interval between the last negative tuberculin test and the first positive tuberculin test. To estimate time of disease onset, we sampled between the last disease negative screening and the first disease positive screening using a gamma distribution from time of tuberculin conversion informed by Poulsen (1957) (33). The population at risk was adjusted to remove individuals with incident disease at the appropriate time while also reflecting loss to follow-up as reported by each study. Data adjustments are discussed in greater detail in Supplemental Information Data Adjustments.

Data to inform transitions between minimal, subclinical, and clinical disease states were extracted from the same systematic review of TB natural history (28), as described in detail elsewhere (29).

### Model development

We expanded a model of the spectrum of TB disease (29) to incorporate *Mtb* infection. Model development relied on an iterative process through which we examined model structures with different pathways from infection to disease states to identify a structure that accurately reflected synthesised data with as simple a structure as possible. The model development process is discussed in Supplemental Information Model Development.

### Model calibration

Using a Bayesian approach, we calibrated the final model to data on progression following *Mtb* infection, as well as data on transitions between minimal, subclinical, and clinical TB disease states (29), mortality (34), duration of infectious TB (35), and prevalence ratios of minimal to infectious TB (18) and subclinical to clinical TB (14). Data were weighted relative to the cohort size, regardless of the number of data points provided (29). All transition rates were assigned uninformed uniform priors [U(0.00,3.00) per year] (29), with the exception of mortality rate [N(0.39,0.03) per year] (34). Informative priors were assigned to the duration of infectious TB [N(2.00,0.50) years] (35), the ratio of minimal to infectious TB prevalence [N(2.50,0.50)] (18), and the ratio of subclinical to clinical TB prevalence [N(1.00,0.25)] (14), as described elsewhere (29). Median values and 95% confidence intervals (CIs) for prior distributions are shown in Figure 1.

**Figure 1:**
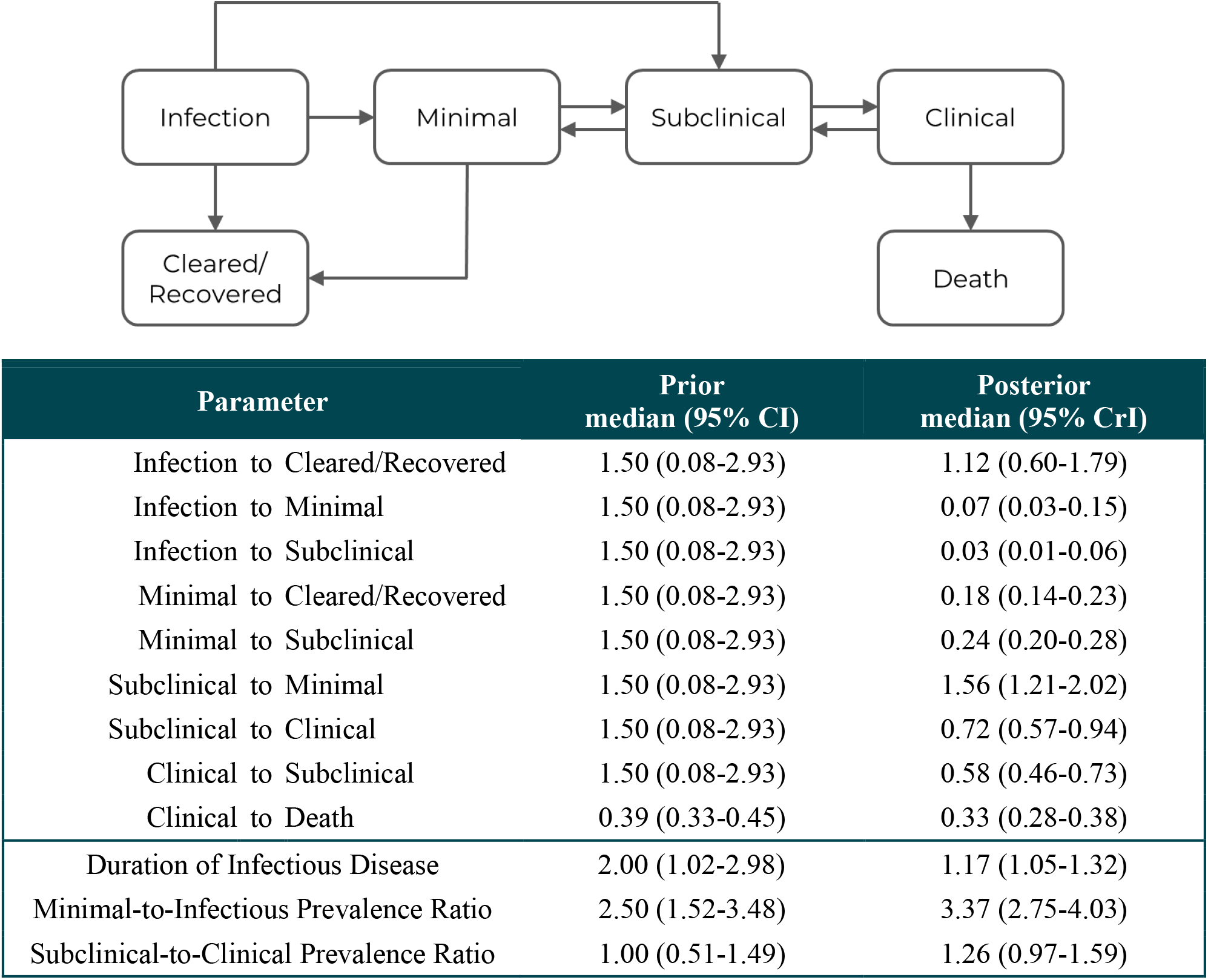
Model structure and prior and posterior parameters showing pathways and progression following *Mtb* infection across the spectrum of TB

Posterior estimates were calculated using a sequential Markov chain Monte-Carlo (MCMC) algorithm in LibBi (36) via R statistical software (37) with RBi (38) and rbi.helpers (39) packages. An adaptive process was used to run repeated chains of 1,000 iterations, adapting parameter values until the acceptance rate was between 20% and 30%. A further 150,000 iterations were then run and the initial adaptive iterations were discarded as burn-in. Convergence of the final 10,000 iterations was assessed visually. Posterior parameter estimates are presented as median values and credible intervals (CrIs).

### Progression following infection

Incidence estimates for minimal, subclinical, and clinical TB were generated using posterior parameter estimates in abbreviated models in which individuals exited the model after reaching the disease state of interest (Supplemental Information Figure 2). As individuals can and do undulate between disease states, defining incidence is not straightforward. Here we counted individuals as incident for each state on their first time entering that state. We report median and 95% uncertainty intervals (UIs) for annual and cumulative incidence over a 10-year period following *Mtb* infection.

### Pathways following infection

To quantify pathways following *Mtb* infection, we simulated 1,000 cohorts of 100,000 individuals over a 10-year period from the time of *Mtb* infection with transitions at monthly intervals. Parameters were sampled independently at the beginning of each simulation for each cohort to capture uncertainty.

Pathways through model states over a 10-year period following *Mtb* infection are reported. Individuals were considered to have experienced “undulation” if they progressed to any disease state, regressed from that disease state, and then progressed again to the same or a more advanced disease state (i.e., changed trajectory at least twice). Median and 95% UIs are reported.

Monthly transitions were also categorised to report a dominant disease state for each individual for each 12-month period following *Mtb* infection. If, during a 12-month period, an individual spent nine or more months in a single state, that state is reported for the 12-month period. If, during a 12-month period, an individual spent less than nine months in a single state or transitioned between states three or more times, the state is reported as “transitional” for the 12-month period (29).

### Role of the funding source

The funders had no role in study design, data collection and analysis, decision to publish, or preparation of the manuscript.

## Results

### Data synthesis

To inform progression from *Mtb* infection to TB disease, we screened 54 studies of TB natural history: 49 from the systematic review (28) and five from other studies (11, 32). We reviewed the full-text of 18 potentially relevant studies, excluded 15 studies (13, 33, 40-58), and identified three studies for inclusion (35, 59-62). Excluded studies are described in Supplemental Information Data Synthesis.

Two studies followed individuals from tuberculin conversion to minimal TB disease. Daniels (1944) (59) describes follow-up of 722 tuberculin-negative nursing students who were enrolled between 1935 and 1943 in London, UK. Tuberculin conversion was detected by annual screening in 248 participants, who were then followed annually for up to five years. Twenty-seven individuals developed TB disease, defined via radiography per Prophit committee standards (59). Madsen (1942) (60) describes follow-up of 1,099 tuberculin-negative medical & high school students enrolled between 1934 and 1936 in Copenhagen, Denmark. Tuberculin conversion was detected by approximately annual screening in 208 participants, who were then followed approximately annually for up to five years. Fifty-two individuals developed TB disease, defined as demonstrable chest x-ray changes following tuberculin conversion (60).

One study followed individuals from tuberculin conversion to infectious TB disease. The National Tuberculosis Institute describes a longitudinal study of TB natural history conducted between 1961 and 1968 in Bangalore, India (35, 61, 62). Four surveys were conducted, with 18-month intervals between the first and second and second and third and a 24-month interval between the third and fourth. Tuberculin testing was performed at each survey for previously tuberculin-negative individuals; disease screening was performed for previously tuberculin-positive individuals. Tuberculin conversion was detected in 1,538 participants. Forty-two individuals developed TB disease, defined as chest x-ray positive and sputum culture-positive, regardless of symptoms (35, 61, 62). Data from this study are included due to the limited evidence available elsewhere, with recognition that investigators’ decision to withhold effective TB treatment from study participants would now be viewed as highly unethical. See Supplemental Information Data Synthesis for more detail.

Weighted data used in model calibration are shown in Figure 2.

**Figure 2:**
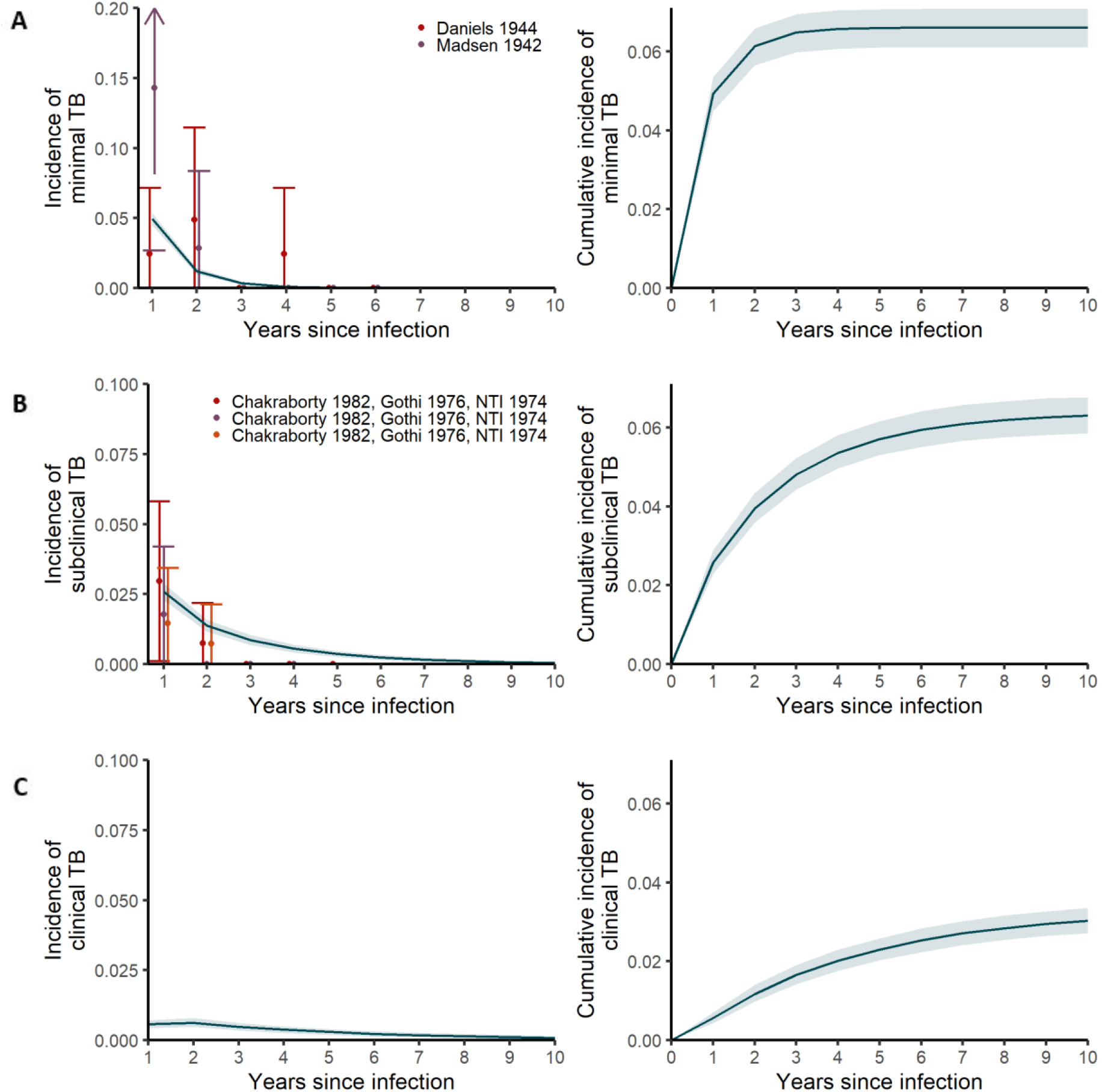
Annual (left) and cumulative (right) incidence for minimal (A), subclinical (B), and clinical (C) TB disease over a 10-year period following *Mtb* infection. Lines and shaded areas show medians and 95% uncertainty intervals for calibrated model. Points and error bars show medians and 95% confidence intervals for calibration targets for minimal and subclinical TB disease.

Data to inform transition rates between minimal, subclinical, and clinical TB disease states, mortality rate, duration of infectious TB, and prevalence ratios for minimal to infectious TB and subclinical to clinical TB are described in detail elsewhere (29).

### Model development

The final model structure reflects heterogeneous progression following *Mtb* infection, with pathways for progression from infection to either minimal or subclinical disease, and allows clearance from infection and recovery from minimal disease (Figure *1*). The model allows progression and regression between minimal, subclinical, and clinical disease states, with TB-associated mortality from the clinical state only (29).

### Model calibration

Medians and 95% credible intervals (CrIs) for posterior parameter distributions are shown in Figure 1. Median rates for progression from infection to minimal disease and progression from infection to subclinical disease were 0.07 (95% CrI 0.03-0.15) and 0.03 (95% CrI 0.01-0.06) per year, respectively. The median clearance rate was 1.11 (95% CrI 0.60-1.80) per year from infection, and the median recovery rate was 0.18 (95% CrI 0.14-0.23) per year from minimal disease. Posterior calibrations for progression from infection to minimal disease and from infection to subclinical disease are shown with weighted calibration targets in Figure 2. Posterior calibrations and weighted calibration targets for transitions between minimal, subclinical, and clinical disease states and for remaining targets are shown in Supplemental Information Figure 4 and Supplemental Information Figure 5, respectively.

### Progression following infection

Over a 10-year period following *Mtb* infection, 90.8% (95% UI 90.2-91.3) of simulated individuals cleared infection without progressing to TB disease, with 83.3% (95% UI 82.7-84.1) clearing infection within the first two years (Figure 3).

**Figure 3:**
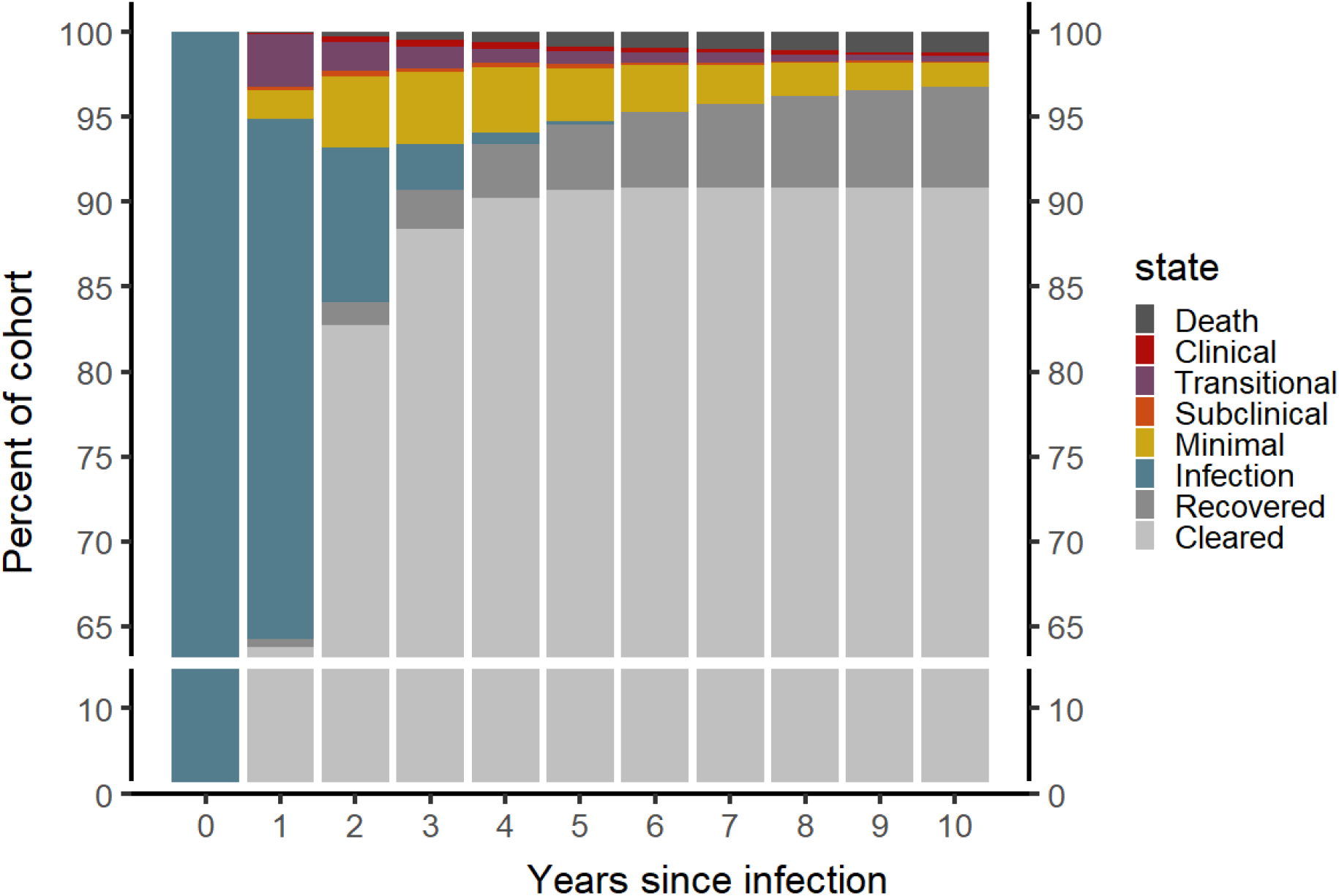
Median distribution of simulated individuals by dominant disease state for each year over 10 years following *Mtb* infection

Cumulative incidence of TB disease was 8.6% (95% UI 7.8-9.4) at two years and 9.3% (95% UI 8.4-10.0) at 10 years. At two years post-infection, cumulative incidence of minimal, subclinical, and clinical TB disease was 6.1% (95% UI 5.7-6.6), 4.0% (95% UI 3.6-4.3), and 1.2% (95% UI 1.0-1.4), respectively. Further incidence over the period from three to 10 years gave a ten-year cumulative incidence of 6.6% (95% UI 6.1-7.1), 6.3 (95% UI 5.9-6.8), and 3.0 (95% UI 2.7-3.4), respectively, for minimal, subclinical, and clinical disease. Annual and cumulative incidence of minimal, subclinical, and clinical TB disease over a 10-year period following *Mtb* infection are shown in Figure 2.

### Pathways following infection

Of the simulated individuals who developed TB disease, 71.6% (95% UI 68.5-74.6) progressed from infection to minimal disease, while 28.4% (95% UI 25.4-31.5) progressed directly from infection to subclinical disease (Figure 4). 31.9% (95% UI 28.9-34.9) of simulated individuals developed minimal disease and did not progress to infectious disease within 10 years of infection. In total, 68.1% (95% UI 65.1-71.1) of simulated individuals who progressed to disease developed subclinical disease. 35.3% (95% UI 32.6-38.7) of simulated individuals developed subclinical disease but did not progress further to clinical disease within 10 years of infection, while 32.7% (95% UI 29.7-35.7) progressed to clinical disease. Of the simulated individuals who developed TB disease, 15.3% (95% UI 13.1-17.5) died from TB-associated mortality in this untreated cohort, and 64.1% (95% UI 61.1-67.3) recovered within 10 years of infection.

**Figure 4:**
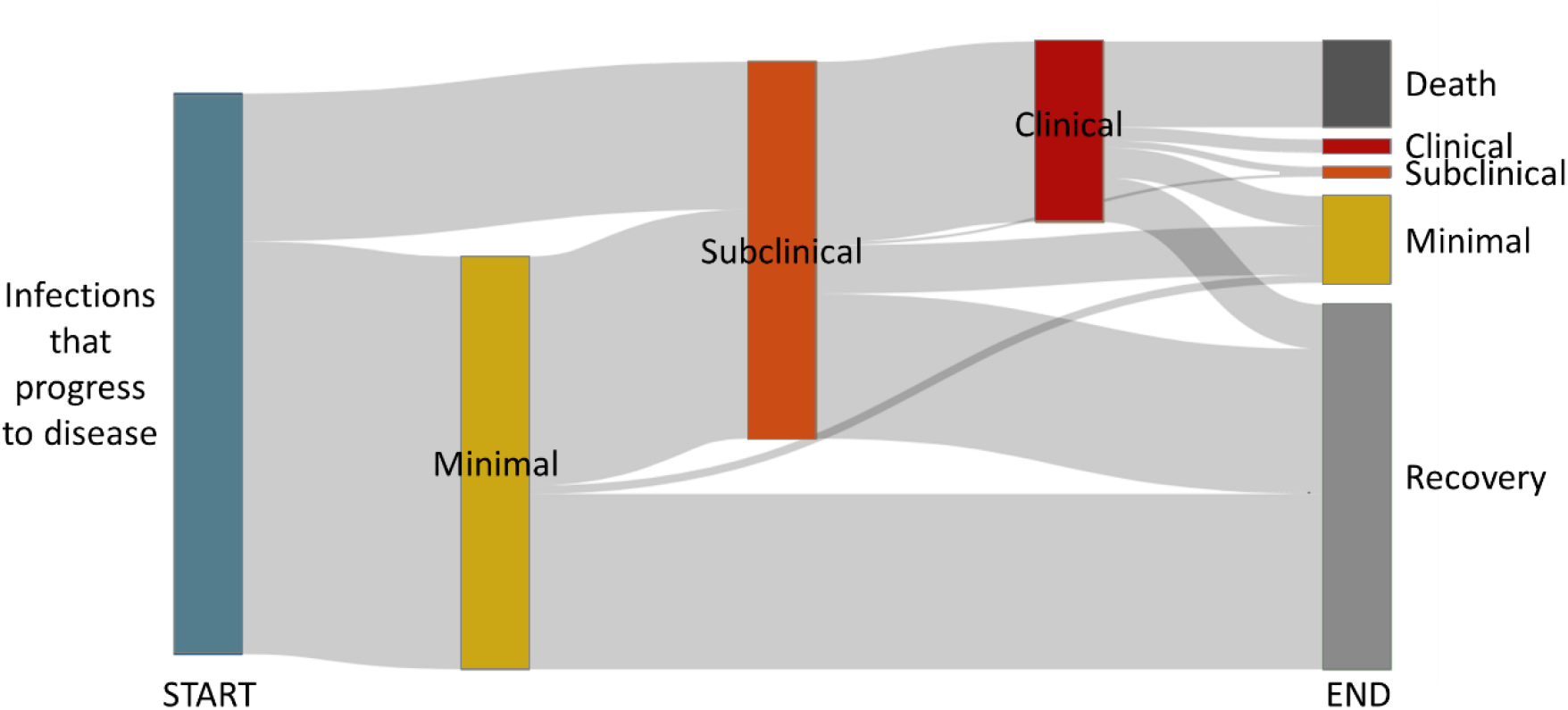
Sankey diagram showing broad pathways over a ten-year period following *Mtb* infection for simulated individuals who developed TB disease. Start represents simulated individuals who progress to TB disease at Year 0. End shows the distribution of those simulated individuals across states at Year 10. Undulation between states is not shown.

Of the simulated individuals who developed TB disease, 56.7% (95% UI 53.6-59.6%) progressed and then regressed towards recovery, either recovering or remaining in a less advanced disease state at the end of the 10-year period. Another 33.3% (95% UI 30.4-36.3%) of the simulated individuals who developed TB disease – 48.8% (95% UI 45.0-52.6) of those who developed infectious disease – undulated between states (i.e., progressed, regressed, then progressed again) in their course of disease. The final 10.0% (95% UI 8.2-12.0%) of simulated individuals who developed TB disease progressed with no regression during the 10-year period following infection.

## Discussion

Our rigorous evaluation of historical data and application within a Bayesian modelling framework allows us to differentiate progression risk and pathways following *Mtb* infection by different disease thresholds across the spectrum of TB disease. Over a ten-year period following *Mtb* infection, one in 10 simulated individuals progressed to TB disease. Of those, two-thirds developed infectious disease, half of which progressed to clinical disease. Timeframes for incidence varied substantially between disease thresholds. While nearly all progression to minimal disease and most progression to subclinical disease occurred within two years of infection, most progression to clinical disease occurred later in the course of disease. Heterogeneous progression pathways from *Mtb* infection were necessary to calibrate the model, and pathways following infection were diverse with half of those who developed infectious disease undulating between disease states.

Our findings show that different thresholds of TB disease have different implications for understanding pathways following *Mtb* infection. The traditional threshold of infectious symptomatic disease recognises only a third of our simulated cohort who progressed beyond *Mtb* infection and after those individuals had progressed through less advanced disease states. A lower threshold acknowledging any infectious disease, regardless of reported symptoms, recognised another third of the cohort in addition to those with infectious, symptomatic disease. This encompassed all those who contribute to *Mtb* transmission (15, 63). As nearly half of individuals who developed subclinical TB went on to develop clinical TB, a threshold of infectious disease has potential individual benefit through the opportunity to avert further morbidity and possible mortality, as well as population benefit from the interruption of further transmission. Yet this threshold still omits a third of individuals who progressed from *Mtb* infection in our study. Though these individuals are considered non-infectious, more than half went on to develop infectious disease, so intervention at this threshold could avert not only more severe morbidity but also truly prevent future transmission.

Our findings emphasise a need for caution and greater attention to definitions when considering broad statements about timelines of disease risk. Our findings are consistent with the lower end of the conventional lifetime risk of progression to TB disease when considering a threshold of minimal or subclinical disease, but our estimated ten-year cumulative incidence of clinical disease is well below that axiom. Our results dispute assertions that the vast majority of infectious, symptomatic TB disease develops less than two years after *Mtb* infection (50). We found that that less than half of progression to clinical disease occurs during this period, though nearly all progression to minimal disease and more than half of progression to subclinical disease occurred within this period.

There is much heterogeneity in pathways following *Mtb* infection. In our model development, we found that data on progression following *Mtb* infection cannot be explained without allowing multiple progression paths, including rapid progression to infectious disease and the opportunity to clear *Mtb* infection. Our simplified pathways reflecting progression to minimal disease and progression to subclinical disease may be considered in line with traditional considerations of “rapid” and “slow” progression following infection. The proportion of self-clearance in our model reflects the inclusion of clearance as an output and the indirect influence of data used for calibration; no informative prior was assigned for the rate of self-clearance. The resulting median 90.8% self-clearance in our simulated cohort is consistent with data-driven estimates from others (27). We have also highlighted that complex pathways through the spectrum of TB disease are common. Pathways reflecting only progression through the course of disease were rare, occurring for only a tenth of individuals who developed disease. More than half of individuals experienced at least some regression towards recovery in their course of disease, while half of those who developed infectious disease undulated between disease states. Transitional periods were also common, particularly in the first years after infection.

Our evaluation of potential data sources to inform this work was rigorous. Although data points are sparce, we rely on three studies which documented timed tuberculin conversions for 2000 individuals (as a proxy for *Mtb* infection) and approximate time of disease onset, as well as information on the population at risk which allowed us to calculate incidence risks over time. This methodology overcomes limitations of other data frequently cited sources (12, 13) that, most often, do not report timing of tuberculin conversion, but may also have unclear disease endpoints or lack data on the underlying population of tuberculin converters at risk of developing disease. Pairing historic (pre-chemotherapy) cohort data on transitions within the spectrum of infection and disease with contemporary (post-chemotherapy) data on the distribution of prevalence across the disease spectrum is a particular strength of our model.

Our work has several limitations. We focus solely on pulmonary TB and cannot comment on extrapulmonary disease. The model is calibrated to separate data for each transition rather than data following trajectories across the spectrum of disease. As such, each transition is independent of any prior disease state or pathway, while in reality previous disease state episodes likely have some influence on future pathways. Our assessment of undulation focuses on progression and regression between states and therefore ignores fluctuations that do not result in transitions between defined states. We also do not consider risk factors for progression from *Mtb* infection, such as age, sex, or HIV infection. Further research is needed to examine whether our findings are consistent across these sub-groups.

Our data-driven modelling work quantifies differences in progression across different thresholds of disease following *Mtb* infection and highlights heterogeneous pathways through the spectrum of TB disease. These results update our understanding of progression risks and timelines in line with the spectrum of TB disease to guide more effective prevention, detection, and treatment efforts and ultimately avert morbidity and transmission in order to end TB.

## Data Availability

All data produced in the present study are available upon reasonable request to the authors.

## Funding statement

KCH, JCE, ASR, and RMGJH received funding from the European Research Council (ERC) under the European Union’s Horizon 2020 programme (Starting Grant Action Number 757699). KCH and ASR are also supported by the UK FCDO (Leaving no-one behind: transforming gendered pathways to health for TB). HE acknowledges support from MRC (MR/V00476X/1). This research has been partially funded by UK aid from the UK government (to KCH and ASR); however the views expressed do not necessarily reflect the UK government’s official policies.

## Notes

### Competing Interest Statement

The authors have declared no competing interest.

