## Supplemental Information for "Re-evaluating progression and pathways following *Mycobacteria tuberculosis* infection within the spectrum of tuberculosis disease"

### I. *Data Synthesis*

The 15 studies excluded after full text review are reported in Supplemental Table 1 with the reason for exclusion for each.

Supplemental Table 1: Reasons for exclusion of potentially relevant studies

| Study | Reason for exclusion |
|---|---|
| Beeuwkes H, Hahn RG, Putnam P. A Survey of Persons Exposed to Tuberculosis in the Household1, The Necessity for Prolonged Observation of Contacts. American Review of Tuberculosis. 1942;45(2):165-93. | Time to disease onset is measured from contact or exposure rather than known tuberculin conversion. |
| Borgen L, Meyer SN, Refsum E. Mass photofluorography, tuberculin testing, and BCG vaccination in the district of Aker (Norway) 1947-49. Acta Tuberculosea Scandinavica. 1951;25(4):327-55.<br>Borgen L, Meyer SN. Mass investigation by photofluorography: An illustration of the value of the method in combating tuberculosis. Acta tuberculosea et pneumologica Scandinavica. 1951;25:288-302. | Time of tuberculin conversion is not reported; disease onset is reported by initial tuberculin test results. |
| Ferebee SH, Mount FW, Murray FJ, Livesay VT. A controlled trial of isoniazid prophylaxis in mental institutions. American Review of Respiratory Disease. 1963;88(2):161-75. | Time of tuberculin conversion is not reported for most participants. Two participants are reported to have converted within 12 months of enrolment, but time to disease onset for these individuals is not reported. |
| Ferebee SH, Mount FW. Tuberculosis morbidity in a controlled trial of the prophylactic use of isoniazid among household contacts. American Review of Respiratory Disease. 1962;85(4):490-510. | Time of tuberculin conversion is not reported. |
| Frimodt-Møller J, Thomas J, Parthasarathy R. Observations on the protective effect of BCG vaccination in a South Indian rural population. Bulletin of the World Health Organization. 1964;30(4):545.<br>Frimodt-Møller J. A community-wide tuberculosis survey in a South Indian rural population, 1950-55. Bulletin of the World Health Organization. 1960;22(1-2):61. | Time of tuberculin conversion is not reported; disease onset is reported by initial tuberculin test results. |
| Gedde-Dahl T. Tuberculous infection in the light of tuberculin matriculation. American Journal of Epidemiology. 1952;56(2):139-214. | Intervals between tuberculin testing are unknown (performed at "suitable intervals"), and intervals between chest x-ray following tuberculin conversion are inconsistent. |
| Hart PA. BCG and vole bacillus vaccines in the prevention of tuberculosis in adolescence and early adult life. Third report to the Medical Research Council by their Tuberculosis | Time of tuberculin conversion is not reported; disease onset is reported by initial tuberculin test results. |

| | |
|---|---|
| Vaccines Clinical Trials Committee. BMJ. 1963;2:973-8.<br>Hart PA. BCG and vole bacillus vaccines in the prevention of tuberculosis in adolescents. First (Progress) report to the Medical Research Council by their tuberculosis vaccines clinical trials committee. British Medical Journal. 1956:413-27.<br>Hart PA. BCG and vole bacillus vaccines in the prevention of tuberculosis in adolescents. Second report to the Medical Research Council by their Tuberculosis Vaccines Clinical Trials Committee. BMJ. 1959;2:379-96. | |
| Israel HL, Hetherington H, Ord JG. A study of tuberculosis among students of nursing. Journal of the American Medical Association. 1941;117(10):839-44. | Disease onset is not reported by tuberculin status. |
| Okada K, Onozaki I, Yamada N, Yoshiyama T, Miura T, Saint S, et al. Epidemiological impact of mass tuberculosis screening: a 2-year follow-up after a national prevalence survey. The International journal of tuberculosis and lung disease. 2012;16(12):1619-24. | Time of tuberculin conversion is not reported. |
| Poulsen A. Some clinical features of tuberculosis. 1. Incubation period. Acta tuberculosea Scandinavica. 1950;24(3-4):311-46.<br>Poulsen A. Some clinical features of tuberculosis. Acta tuberculosea Scandinavica. 1957;33(1-2):37. | Intervals between tuberculin testing are uncertain and inconsistent across the population ("the majority of the population has been submitted to systematic tuberculin tests… at intervals of one or two years") and not all "converters" had prior evidence of a negative tuberculin reaction. |
| Rubinstein H, Kotschnowa I. Beginn und entwicklung der lungentuberkulose beim erwachsenen. Acta Medica URSS. 1940;3(3):250-65. | Time of tuberculin conversion is not reported; disease onset is reported by initial chest x-ray results. |
| Sikand B, Narain R, Mathur G. Incidence of TB as Judged by Re-surveys A Study of Delhi Police By. 1959. | Time of tuberculin conversion is not reported; tuberculin testing was only performed at enrolment. |
| Stýblo K, Dankova D, Drapela J, Galliova J, Ježek Z, Křivánek J, et al. Epidemiological and clinical study of tuberculosis in the district of Kolin, Czechoslovakia: report for the first 4 years of the study (1961-64). Bulletin of the World Health Organization. 1967;37(6):819. | Time of tuberculin conversion is not reported; disease onset is reported by initial chest x-ray results. |
| Sutherland I. Recent studies in the epidemiology of tuberculosis, based on the risk of being infected with tubercle bacilli. Advances in tuberculosis research Fortschritte der Tuberkuloseforschung Progres de l'exploration de la tuberculose. 1976;19:1-63 | Indirect population-level estimates of progression |
| Wallgren A. The time-table of tuberculosis. Tubercle. 1948;29(11):245-51. | No description of the study methodology underlying reported time between exposure and disease onset could be found. |

Data from the longitudinal study of TB natural history conducted by the National Tuberculosis Institute in Bangalore, India, are included as evidence of progression following *Mtb* infection. Authors of this study, conducted between 1961 and 1968, state that "no organized antituberculosis treatment was available to the people of the area during the entire study period" (1). However, they also report that "[d]uring the second survey and the early part of the third, with a view to obtaining better cooperation, one month's supply of isoniazid tablets was issued to persons in whom pulmonary tuberculosis was diagnosed" (1). This limited course of isoniazid would have had little effect on disease progression and may have contributed to the emergence of drug resistant strains of *Mtb*. Authors' acknowledgement of this limited provision implies that investigators had knowledge of and access to isoniazid, yet it appears no efforts were made to provide an effective regimen of the drug to study participants outside the scope of improving "cooperation". We have decided to include data from this research, which would now be considered highly unethical, because we were unable to identify any other data sources that could provide comparable data, and we feel not using the data would make the burden placed on study participants even more onerous.

*II. Data Adjustments*

After extracting data from three included studies describing progression from *Mtb* infection to TB disease, we adjusted those data to reflect uncertainty in the time of tuberculin conversion and disease onset, recognising that neither infection nor disease onset occur at the point those developments are detected in these studies. For studies reporting progression from TST conversion to minimal disease, we first reduced the number of reported cases by 25% to acknowledge that some pathological anomalies detected by chest x-ray may not have been attributable to TB disease (2).

For all studies, to estimate time of infection, we sampled from a uniform distribution over the interval between the last negative tuberculin test and the first positive tuberculin test.

For all studies, to estimate time of disease onset, we sampled between the last disease negative screening and the first disease positive screening using a gamma distribution. The gamma distribution [$\Gamma(2.2620958,0.9581921)$] was informed by data from Poulsen (1957) (3) on the cumulative proportion of incident cases of TB disease over time, shown in Supplemental Table 2. The gamma distribution was aligned with the previously sampled time of tuberculin conversion and truncated to sample within the interval between the last negative disease screen and the first positive disease screen.

Supplemental Table 2: Cumulative proportion of incident cases of TB disease by years since infection per Poulsen

| **Years since infection** | **Cumulative proportion of incident cases** |
|---|---|
| 1 | 0.17 |

| | |
|---|---|
| 2 | 0.52 |
| 3 | 0.70 |
| 4 | 0.87 |
| 5 | 0.93 |
| 6 | 0.97 |
| 8 | 0.98 |
| 10 | 1.00 |

We adjusted the number of tuberculin converters at risk of disease to remove individuals with incident disease at the appropriate time while also reflecting loss to follow-up as reported by each study. We note that no loss to follow-up was reported in manuscripts describing the National Tuberculosis Institute study in Bangalore, India, which risks underestimating progression to subclinical disease in later years.

*III. Model Development*

The model structure relating *Mtb* infection to TB disease states was developed through an iterative process. In total, we examined 4 potential model structures defining progression from *Mtb* infection in different ways (Supplemental Figure 1). Model A allowed only progression from infection to minimal disease. Model B allowed direct progression from infection to subclinical disease in addition to progression from infection to minimal disease. Model C allowed progression to subclinical disease, progression to minimal disease, and indirect progression to minimal disease via an intermediate state. Model D allowed direct progression from infection to clinical disease, in addition to progression from infection to minimal disease and progression from infection to subclinical disease.

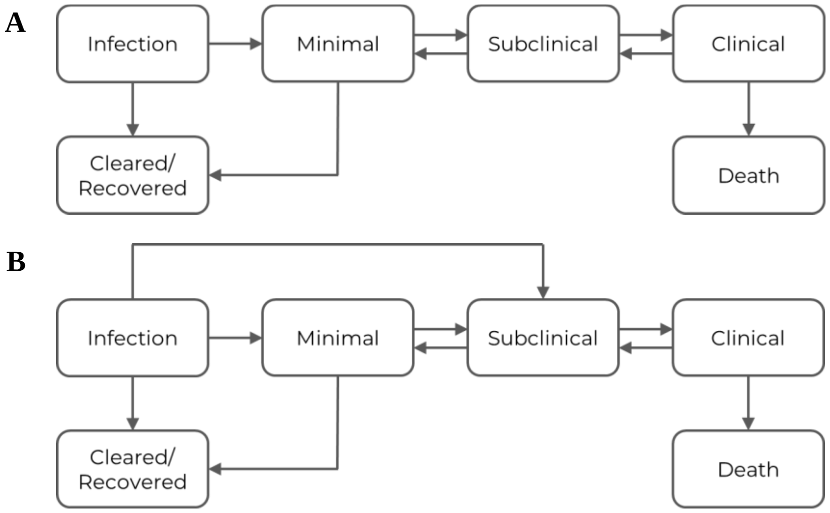

**C**

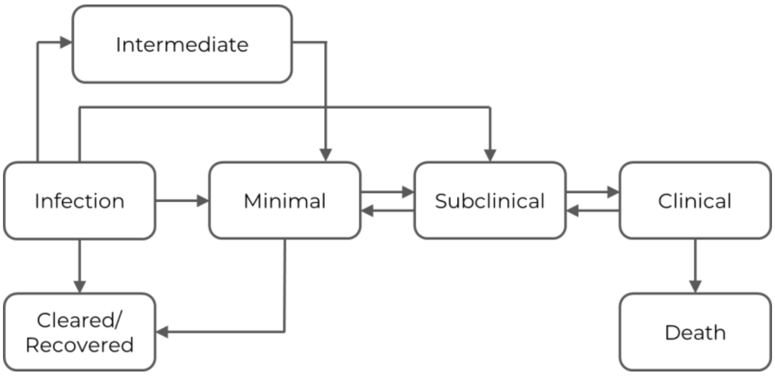


**D**

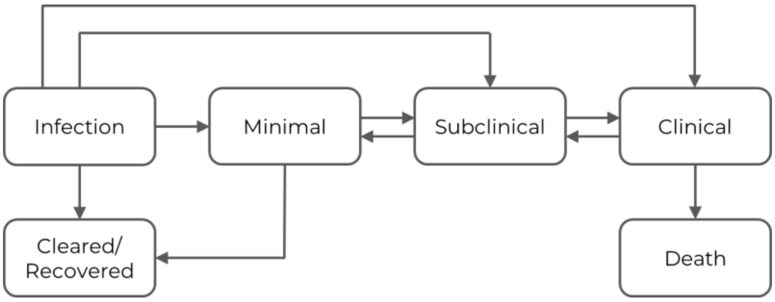


Supplemental Figure 1: Potential model structures examined during the model development process

Each model structure was calibrated following methods described in the main text Methods and Supplemental Information Model Calibration. Models were evaluated based on deviance information criteria (DIC) and visual inspection of posterior calibrations to progression from infection to minimal disease and from infection to subclinical disease. Calibrations for each model are shown in Supplemental Table 3.

Supplemental Table 3: Model development calibrations

| | Calibration to minimal disease | Calibration to subclinical disease | DIC |
|---|---|---|---|
| A | 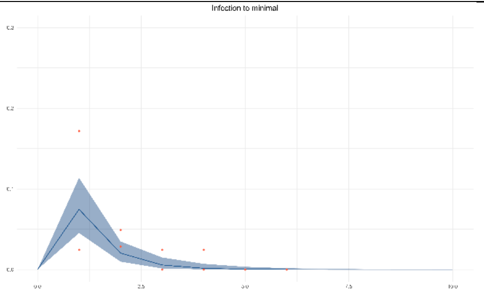 | 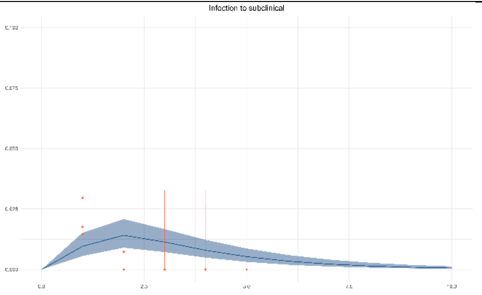 | 685.8 |
| B | 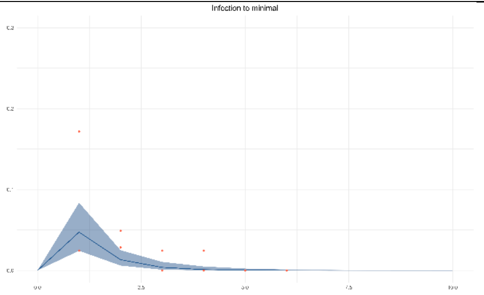 | 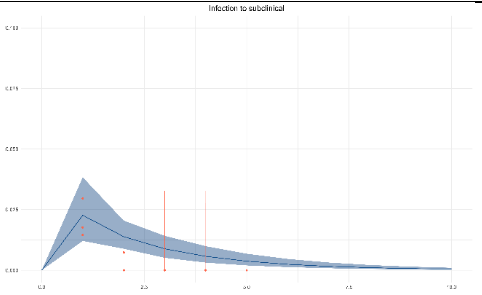 | 678.2 |

| | | | |
|---|---|---|---|
| C | 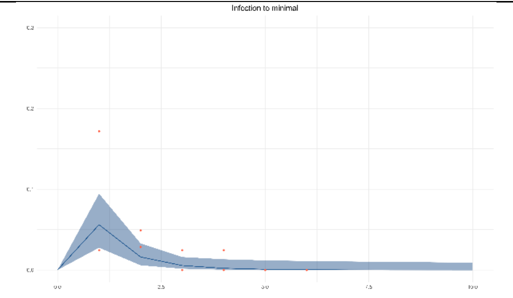  | 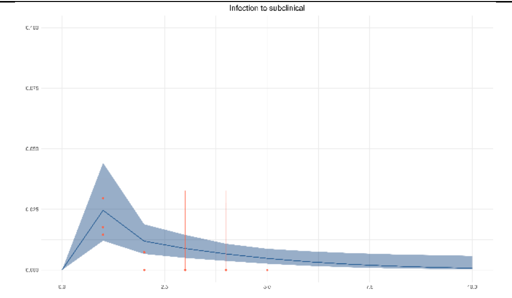  | 685.1 |
| D | 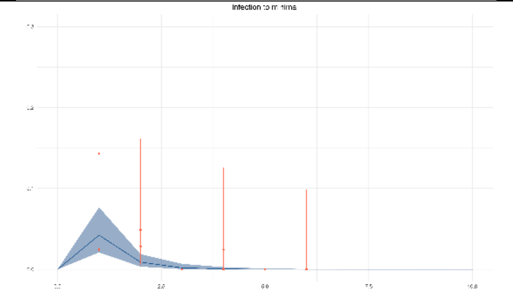 | 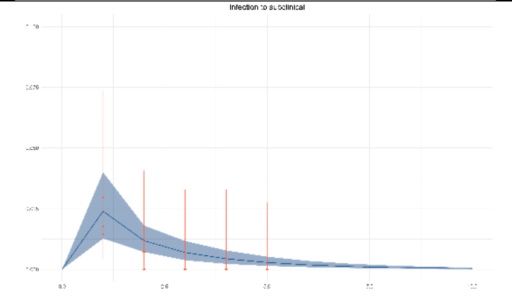 | 671.2 |

For Model A, the calibrated incidence of minimal disease was considered reasonable, but the shape of the calibrated incidence of subclinical disease deviated from the shape of the data for this calibration target. For Model B, the shape of the calibrated incidence of subclinical disease was improved, relative to Model A, as was the overall fit. For Model C, there was little visual difference in the calibrated incidence of minimal and subclinical disease, relative to Model B, and the overall fit was comparable to that in Model A. For Model D, there was little visual difference in the calibrated incidence of minimal and subclinical disease, relative to Model B or Model C, though the overall fit was improved. The improved overall fit in Model D was not considered sufficient to compensate for the added model complexity, so Model B was selected for the main model analysis.

*IV. Model Calibration*

The equations used to describe the main model are as follows:

$$\frac{dI}{dt} = -(infclear + infmin + infsub) * I$$

$$\frac{dM}{dt} = infmin * I - (minclear + minsub) * M + submin * S$$

$$\frac{dS}{dt} = infsub * I + minsub * M - (submin + subclin) * S + clinsub * C$$

$$\frac{dC}{dt} = subclin * S - (clinsub + clinmort) * C$$

where I, M, S, and C refer to infection, minimal, subclinical, and clinical states, respectively, and remaining terms represent transition rates with *infclear* from infection to cleared/recovered, *infmin* from infection to minimal, *minclear* from minimal to cleared/recovered, *minsub* from minimal to subclinical, *submin* from subclinical to minimal, *infsub* from infection to subclinical, *subclin* from subclinical to clinical, *clinsub* from clinical to subclinical, and *clinmort* from clinical to death.

Appropriate sub-structures from this model were used in the calibration process to align with the data informing calibration targets for different transitions (2). Model structures used to calculate incidence of minimal, subclinical, and clinical disease are shown in Supplemental Figure 2.

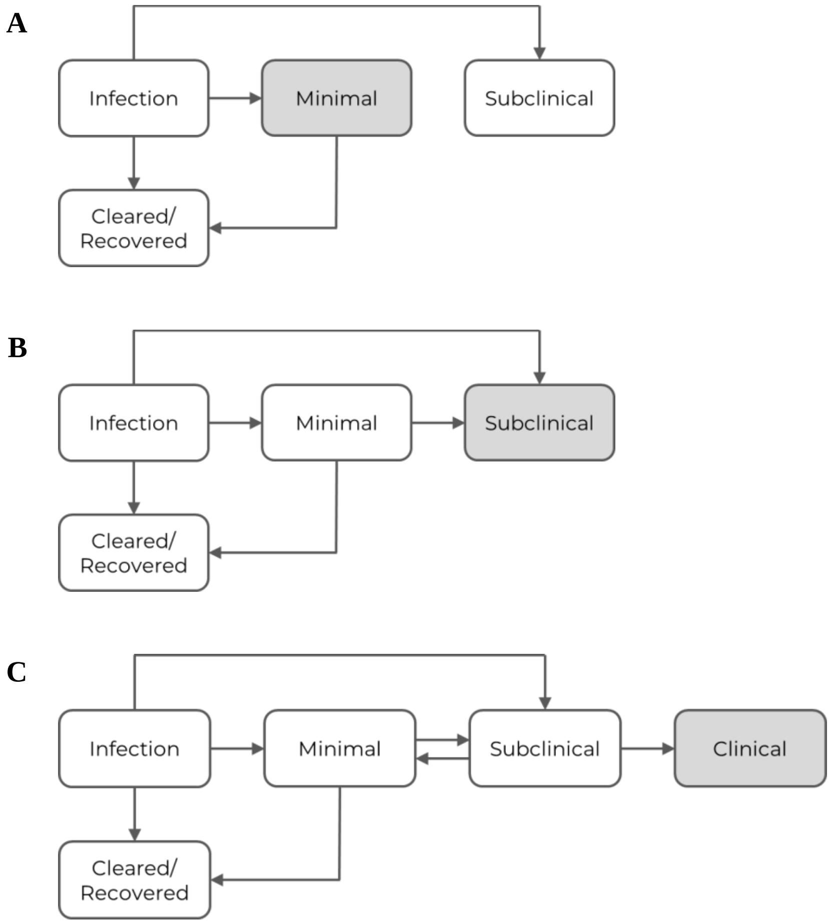


Supplemental Figure 2: Model structure to quantify incidence of minimal (A), subclinical (B), and clinical (C) TB disease

Trace plots for each parameter over the 150,000 iterations of the calibrated model are shown in Supplemental Figure 3.

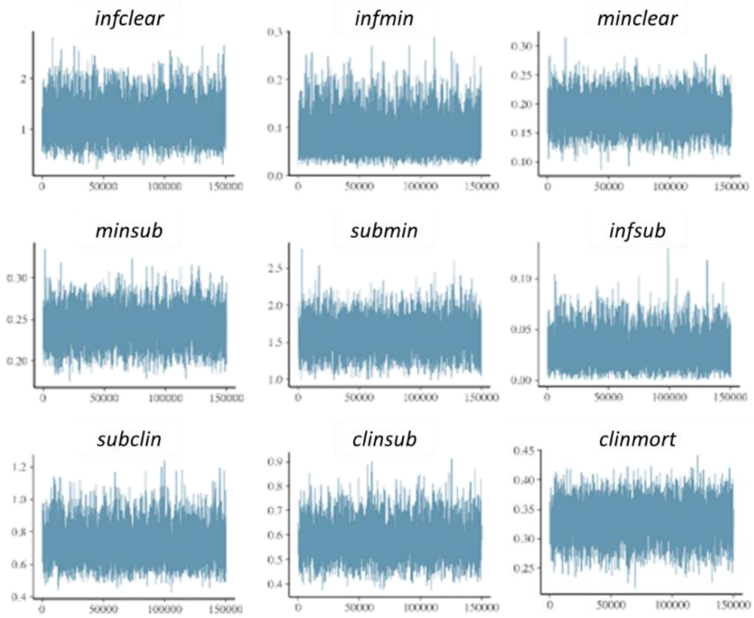


Supplemental Figure 3: Trace plots

Posterior calibrations and weighted calibration targets for transitions between minimal, subclinical, and clinical disease states are shown in Supplemental Figure 4; calibrations and targets for additional contemporary data points are shown in Supplemental Figure 5.

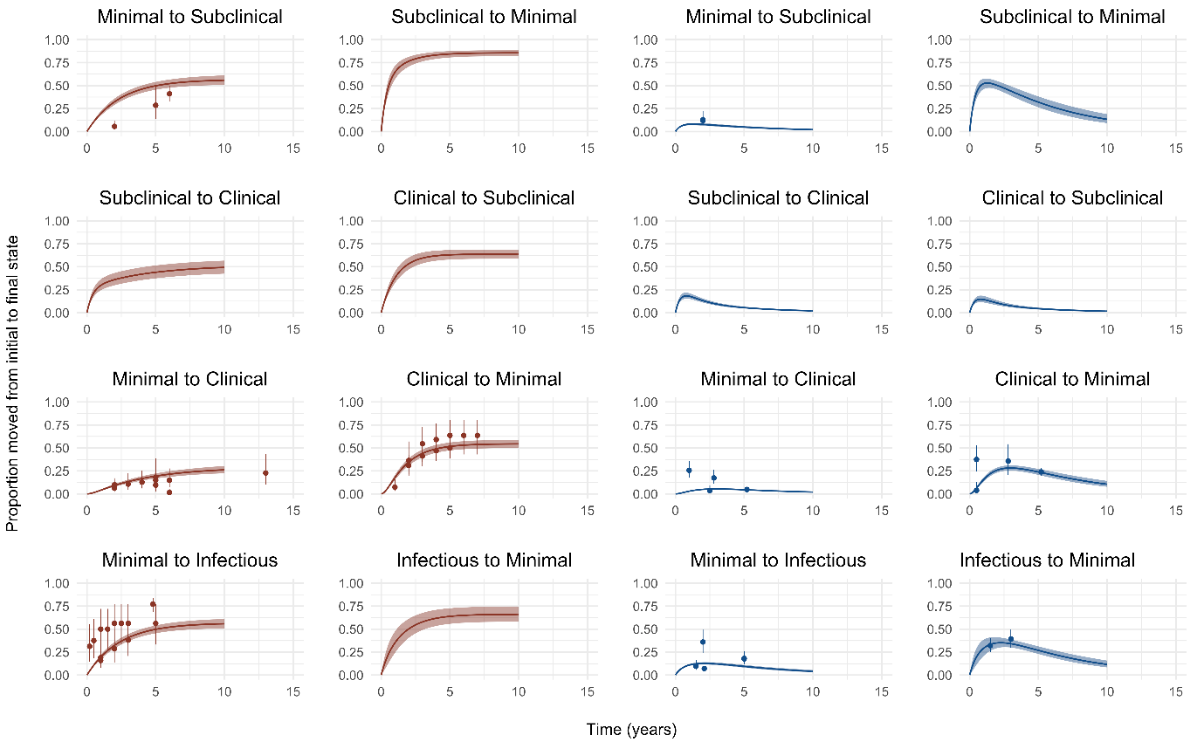


Supplemental Figure 4: Model calibration for transitions between minimal, subclinical, and clinical disease states

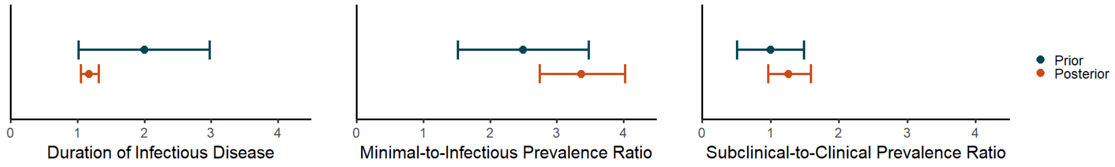


Supplemental Figure 5: Model calibration for additional contemporary data points

### *V. Sensitivity Analysis*

We conducted sensitivity analyses to explore uncertainty in the disease states represented by NTI data due to the lack of reporting of any clinical signs or symptoms. We examined three scenarios with different degrees of clinical representation among cases, classifying cases as either 0%, 50%, or 100% clinical. Calibrations are shown in Supplemental Figure 6.

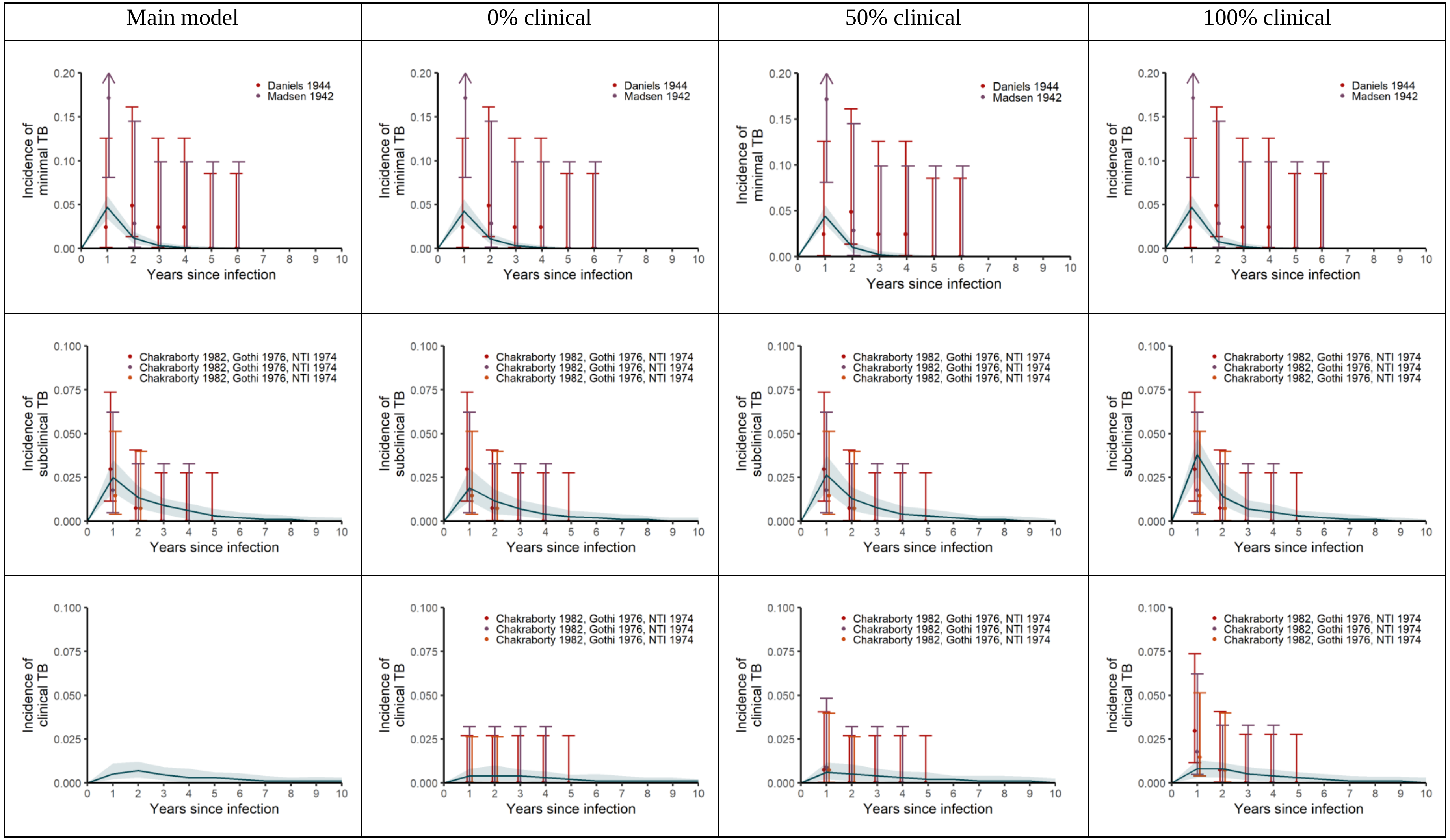


Supplemental Figure 6: Sensitivity analysis calibrations for different classifications of NTI data

*VI. References*


1. National Tuberculosis Institute, Tuberculosis in a rural population of South India: a five-year epidemiological study. *Bulletin of the World Health Organization* **51**, 473 (1974).

2. A. S. Richards *et al.*, The natural history of TB disease-a synthesis of data to quantify progression and regression across the spectrum. *medRxiv* (2021).

3. A. Poulsen, Some clinical features of tuberculosis. *Acta Tuberculosea Scandinavica* **33**, 37 (1957).
